# TwoStepCisMR: A novel method and R package for attenuating bias in *cis*-MR

**DOI:** 10.1101/2022.06.13.22276262

**Authors:** Benjamin Woolf, Dipender Gill

## Abstract

Mendelian randomisation (MR) is an increasingly popular method for strengthening causal inference in epidemiological studies. cis-MR in particular uses genetic variants in the gene region for a genetic proxy of a drug target to provide quasi-experimental evidence for drug efficacy. A major problem for this framework is when the causal variant is correlated to another variant which effects the outcome of interest (confounding through linkage disequilibrium). Methods for correcting bias such as multivariable MR struggle in a cis setting because of the high correlation among genetic variants. Here, we therefore present an alternative method for attenuating bias which does not suffer from this problem. We have additionally developed a simple R package to facilitate the implementation of the method.

---

Mendelian randomisation (MR) is the use of genetic variants in instrumental variables analyses.(1,2) Multivariable Mendelian randomisation (MVMR) is an extension of MR to account for multiple exposures. MVMR was developed for mediation analysis, but has more recently been used to attenuate potential biases related to pleiotropic associations of genetic variants.(3) An MR analysis is ‘*cis*’ when genetic variants close to a target gene region are selected as instruments. A major source of bias in *cis*-MR is confounding by a genetic variant in linkage disequilibrium (LD).(4) Although it is possible to explore this using colocalization, this approach is not able to generate MR estimates that account for any such bias.(5)

An alternative to MVMR for performing mediation analyses is two-step MR (TSMR).(6) TSMR leverages the product & dereference of coefficients methods to perform mediation analyses using MR estimates. TSMR has not, to our knowledge, been used to attenuate bias, but we believe that the product of coefficients method could be used for this purpose in *cis*-MR, which we dub Two-Step cis-MR (TSCMR). Specifically, the crude variant-outcome association can be decomposed into (1) a path from the variant to the outcome via the exposure, and (2) a path from the variant to the outcome via the confounder (Figures 1 and 2). It follows that if we can estimate the second path, we could derive an unbiased variant-outcome association estimate by subtracting the effect through the second path from the total effect of the variant on the outcome. This adjusted variant-outcome association could then be used in a *cis*-MR analysis. Intuitively, because we would have removed the effect of any pathway that goes through the confounder phenotype, the revised MR estimate should be unbiased. The second path is itself the product of the effect of the variant on the confounder and the effect of the confounder on the outcome. The first of these will be directly estimated by a genome-wide association study (GWAS) of the confounder. The second could be estimated through MR of the confounder phenotype on the outcome. The standard error for the variant-outcome association should then be updated using the either the propagation of error method or via bootstrapping (see the Supplementary Methods for more details). The only major difference between TSCMR and TSMR is that the latter uses two MR estimates to calculate the indirect effect of an exposure on an outcome, whereas TSCMR uses one MR estimate, and two variant-phenotype estimates, to estimate the direct effect of the variant on the outcome.

**Figure 1:**
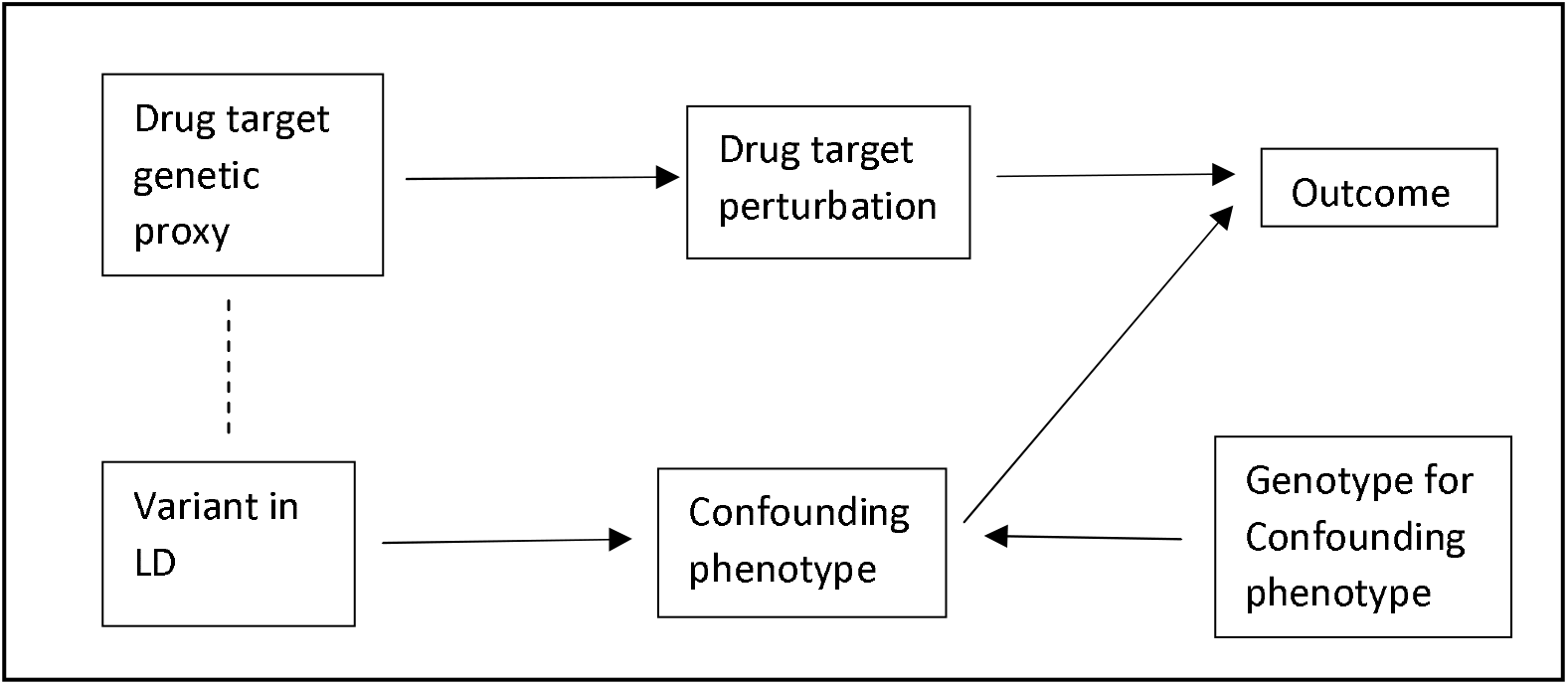
DAG representing confounding by LD in a cis-MR analysis.

**Figure 2:**
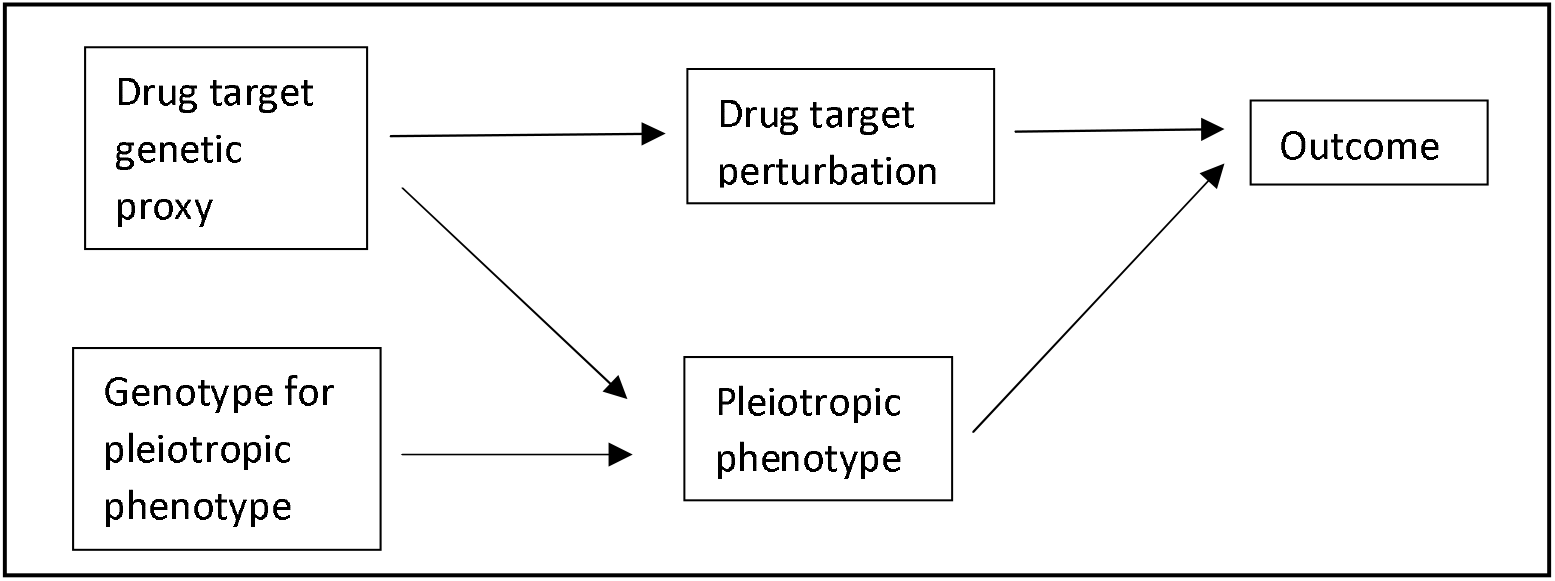
DAG representing bias through pleiotropy in a *cis*-MR analysis.

We used a simulation and an applied example to explore this use of TSCMR further. In brief, our simulation modelled a situation where a second phenotype is either pleiotropic or caused by a variant in perfect LD with the causal variant for the exposure of interest (represented by the Directed Acyclic Graph in Figure 2). We estimated the mean bias and precision of TSCMR compared to traditional *cis*-MR after 100,000 repetitions. Reporting of the methods following the ADEMP guidelines can be found in the Supplementary Methods.(7) A formal proof of the validity of using two-step methods for mediation analyses when using an MR framework can be found elsewhere.(8)

As an applied example, we then looked at the effect of interleukin 6 receptor (IL6R) signalling on coronary artery disease (CAD) risk, and whether the genetic association was confounded by a variant in LD that associated with type-2 diabetes (T2D) risk. Because T2D can also cause CAD, there is potential for confounding by LD. Full methods reporting based on the STROBE-MR guidelines are in the Supplementary Methods.(9)

In the simulation, the crude *cis*-MR estimate was highly biased, with an average inflation 38% when compared to the true effect. TSCMR completely attenuated the bias in the crude estimate with only a moderate reduction in precision (Table 1). We therefore believe this result supports that TSCMR can be used to attenuate bias.

**Table 1:**
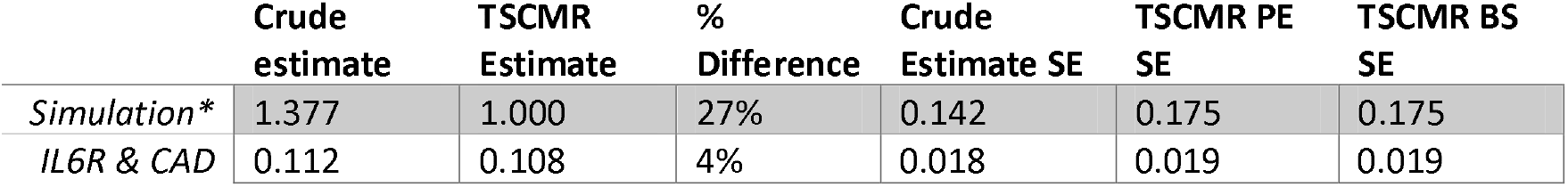
Results of simulation and applied example. Crude = MR estimates using the unadjusted variant-outcome association(s), TSCMR = MR estimates after adjusting the variant-outcome association(s) using TSCMR. SE = Standard error. PE = Propagation of Error. BS = Bootstrapped. The % Difference was defined as the % deflation in the TSCMR estimate compared to the crude estimate. *The estimates for the simulation are the mean estimate after 100,000 repetitions. The true casual effect of the exposure on the outcome in the simulation was 1.000, therefore the mean bias in the effects were 0.909 and 0.029 for the crude and TSCMR models respectively. The Monte-Carlo standard error were < 0.001 for all the estimates other than the bias in the crude effect estimate (Monte Carlo SE = 0.004)

|  | Crude estimate | TSCMR Estimate | % Difference | Crude Estimate SE | TSCMR PE SE | TSCMR BS SE |
| --- | --- | --- | --- | --- | --- | --- |
| *Simulation\** | 1.377 | 1.000 | 27% | 0.142 | 0.175 | 0.175 |
| *IL6R & CAD* | 0.112 | 0.108 | 4% | 0.018 | 0.019 | 0.019 |

We found a 4% deflation in the MR estimate of IL6R signalling on CAD risk after accounting for a confounding path via T2D. This implies the presence of only very moderate degree of confounding by LD in the effect of IL6R signalling on CAD. Since the change in effect is half the size of the standard error, we do not believe that this should be a serious threat to the interval validity of studies exploring the association between IL6R and CAD.(10)

Supplementary Table 1 shows the results of an additional simulation which found that TSCMR is extendable to account for multiple biasing pathways provided that all biasing pathways are independent of each other. Supplementary Box 1 presents a set of limitations to the application of TSCMR. We developed the TwoStepCisMR R package (available from https://github.com/bar-woolf/TwoStepCisMR/wiki) to facilitate the implementation of Two-Step *cis*-MR. We hope that Two-step cis-MR will be a useful sensitivity analyses for MR researchers.

## Data Availability

All the data used in the applied example is available from the MRC-IEU OpenGWAS platform. The R code scripts used in the simulation and applied example are available from https://doi.org/10.17605/OSF.IO/3XG92.

## Declarations

DG is employed part-time by Novo Nordisk. BW declares no conflicts of interest.

## Notes

### Funding Statement

Benjamin Woolf is funded by an Economic and Social Research Council (ESRC) South West Doctoral Training Partnership (SWDTP) 1+3 PhD Studentship Award (ES/P000630/1).

