## Supplementary Methods for "TwoStepCisMR: A novel method and R package for attenuating bias in *cis*-MR"

**Derivation of the standard errors**

Propagation of error

Under the propagation of error rules

$${SE}_{A+B}=\sqrt{{SE}_{A}^{2}+{SE}_{B}^{2}}$$

and

$${SE}_{A*B}= A*B*\sqrt{\left( {\frac{{SE}_{A}}{B_{A}}}^{2}+{\frac{{SE}_{B}}{B_{B}}}^{2} \right)}$$

The estimate we are interested in is:

$$B_{g-o (adjsuted)}= B_{g-o (crude)}- B_{g-c}*B_{c-o}$$

Where B_g-o (adjusted)_ is the adjusted variant-outcome association, B_g-o (crude)_ is the crude variant-outcome association, B_g-c_ is the variant-confounder association, and B_c-o_ is the confounder-outcome association.

It follows that the SE from following the rules of propagation of error rules is:

$$EPSE= \sqrt{{SE}_{g-o (curde)}^{2}+ \left( B_{g-c}*B_{c-o} \right)^{2}*\left( {\frac{{SE}_{g-c}}{B_{g-c}}}^{2}+{\frac{{SE}_{c-o}}{B_{c-o}}}^{2} \right)}$$

Where EPSE is the propagation of error standard error, SE_g-o (crude)_ is the standard error of the crude variant-outcome association, B_g-c_ is the variant-confounder association, and B_c-o_ is the confounder-outcome association, SE_g-c_ is the standard error of the variant-confounder association, and SE_c-o_ is the standard error of the confounder-outcome association.

Bootstrap standard error

Let:

$$D_{g-o (crude)}=N\sim(B_{g-o (crude)}, {{SE}_{g-o (crude)}}^{2})$$

$$D_{g-c}=N\sim(B_{g-c}, {{SE}_{g-c}}^{2})$$

$$D_{c-o}=N\sim(B_{c-o}, {{SE}_{c-o}}^{2})$$

Where, B_g-o (crude)_ is the crude variant-outcome association, SE_g-o (crude)_ is the standard error of the crude variant-outcome association, B_g-c_ is the variant-confounder association, and B_c-o_ is the confounder-outcome association, SE_g-c_ is the standard error of the variant-confounder association, and SE_c-o_ is the standard error of the confounder-outcome association.

Then if we define D’ as:

$$D^{'}=D_{g-o (crude)}-(D_{g-c}* D_{c-o})$$

The bootstrap standard error can then be defined as the standard deviation in D_g-o (crude)_:

$$BSSE=\sqrt{\frac{\sum_{i}^{n} \left( {D'}_{i}- \bar{D'} \right)^{2}}{n}}$$

Where BSSE is the bootstrap standard error, and n is the number of iterations used in the bootstrap.

**Simulation methods**

We report our simulations using the ADEMP (aims, data-generating mechanisms, estimands, methods, and performance measures) approach:(1)

*Aims*: The aim of this simulation was, as a simple proof of concept, to explore if Two-step *cis*-MR can be used attenuate bias in a *cis*-MR analysis.

*Data-generating mechanisms*: The directed acyclic graph (DAG) in Figure 2 was used as the basis for the data generation mechanism for the simulation. The nodes were simulated as standard normal variables (mean = 0, standard deviation [SD] = 1). Each arrow was used to represent a linear causal effect of the node from which the arrow originated on the node to which the arrow was directed, with each beta value being set as 1. The exception to this was the *cis*-variant, which was simulated as a three-level categorical variable with a minor allele frequency of 0.5 (standard deviation = 0.1), and a mean effect size of 0.1 (standard deviation = 0.05). The variant-exposure association, variant-outcome association(s), variant-confounder association(s), were each estimated in independent samples of 200,000 participants. The simulations were repeated 100,000 times.

*Estimands and other targets*: The estimand was the average causal effect of intervening and changing the exposure of each individual from its observed level x by a single unit, or E[Y(X=x)] – E[Y(X=x-1)] using potential outcomes framework.

*Methods*: We compare two methods of estimating the variant-outcome association: 1) the crude association that would be derived from a GWAS, 2) the adjusted association that would be derived by using Two-step *cis*-MR. The confounder-outcome association was estimated as the Wald ratio of association the non-cis genetic risk score for the confounder (GRS) with the outcome divided by the GRS-confounder association. The standard error was estimated as the standard error of the confounder (GRS) with the outcome divided by the absolute value of the GRS-confounder association.

*Performance measure:* The performance measure was the estimate of the casual effect, and the standard error in the estimate of the causal effect for the crude and Two-step cis-MR methods.

**Applied example methods**

Overview

In our applied example we sought to explore the extent to which type 2 diabetes liability confounds the association between IL6R signalling and CAD. We first ran a traditional *cis*-MR of the effect of IL6R signalling (proxied using C-reactive protein [CRP] levels) on CAD risk. We then used Two-step *cis*-MR to adjust the variant-CAD associations for any bias through type 2 diabetes liability. We used variant-type 2 diabetes information extracted form a GWAS, and an MR analysis of the effect of type 2 diabetes liability on CAD risk as the sources of additional information needed to run Two-step *cis*-MR.

Data sources

We used serum C-reactive protein levels as a proxy for IL6R signalling. We extracted CRP GWAS data (OpenGWAS ID: ukb-d-30710_raw) from the Ben Neale lab round 2 analysis.(2) This GWAS included around 469000 males and females of European ancestry. Samples were measured at recruitment. Effect estimates are on the mg/L scale.

We used van der Harst and Verweij (2017)’s GWAS as a source of genotype-CAD associations (OpenGWAS ID: ebi-a-GCST005195).(3) This GWAS was a meta-analysis of the CARDIoGRAMplusC4D and UK Biobank GWASs. Both of these studies are European samples of men and women. This meta-analysis included 122733 cases and 424528 controls. The effect estimates were on the logOR scale. More details can be found in the original manuscript. (3)

Information on variant-(type-2-)diabetes associations were extracted from the FinnGen round 5 GWAS of diabetes (OpenGWAS ID: (finn-b-T2D).(4) This GWAS has information on 29193 cases and 182573 controls, drawn from the Finnish general population. Cases are inferred from medical records. The effect estimates were on the logOR scale.

Instrument construction

We selected genetic instruments which had a genome-wide significant (p < 5 x 10^-8^) association with the exposure (i.e. CRP or type 2 diabetes). We additionally clumped the variants using an r^2^ of 0.1 and KB of 10,000. We used the TwoSampleMR R package to harmonise the two GWASs and to infer which strand was positive. SNPs which were associated with type 2 diabetes but were missing in the CAD GWAS were imputed for the analysis of the effect of diabetes on CAD using an r^2 of 0.8 from the European subsample of the 1000 genomes project.

Statistical methods

The primary MR estimator was the Wald ratio, defined as the variant-outcome association divided by the variant-exposure association. We repeated the analysis using the unadjusted variant-outcome association, and the variant-outcome association estimated using TSCMR.

We used four methods of meta-analysing the SNP specific Wald ratios for the effect of type 2 diabetes liability on CAD and IL6R signalling on CAD: inverse variance weighed (IVW), MR-Egger, weighted median, weighted mode. Unlike IVW, the latter three methods can return the true effect if some of the instruments are invalid, but have reduced power. Weighted mode assumes that the modal effect size is valid estimates of the true effect size, while weighted median assumes that at least half of the SNPs are valid. MR-Egger is unbiased as long as the variant-exposure effect size is independent of the size of any bias (such as a pleiotropic effect).(5) When the Cochrane Q statistic indicated that there was pleiotropy we used the weighted median as the primary estimator.

Assessment of assumptions

Weak instrument bias is inversely proportional to the F-statistic of the variant-exposure association, which we estimated as the square of the variant-exposure association divided by the square of the standard error of this association. We also used the Cochrane’s Q statistic of the Wald ratios as a falsification test for horizontal pleiotropy in the SNP-outcome association.

Software and Preregistration

MR analyses in this paper were run using the TwoSampleMR, R package. All GWAS data was extracted from the MRC-IEU OpenGWAS platform (6).

**Additional simulation methods**

We report our simulations using the ADEMP (aims, data-generating mechanisms, estimands, methods, and performance measures) approach:(1)

*Aims*: The aim of this simulation was to explore if Two-step *cis*-MR can be used attenuate bias in a *cis*-MR analysis when there are multiple known biasing pathways. We explore two settings: 1) when the two pathways are independent, and 2) when they are not independent. For simplicity, in the second setting, we simulated a setting in which two variables on the same pathway were adjusted for.

*Data-generating mechanisms*: The directed acyclic graph (DAG) in Supplementary Figures 1 and 2 were used as the basis for the data generation mechanism for the simulation. As before, this would simulate a setting in which the bias occurs through a pleiotropic SNP or a confounding by a SNP in perfect LD. However, unlike in the pervious setting, both of these data generative models included two variables for adjustment. The parameters were otherwise the same as in the previous simulation. The simulations were again repeated 100,000 times.

*Estimands and other targets*: The estimand was the average causal effect of intervening and changing the exposure of each individual from its observed level x by a single unit, or E[Y(X=x)] – E[Y(X=x-1)] using potential outcomes framework.

*Methods*: We compare two methods of estimating the variant-outcome association: 1) the crude association that would be derived from a GWAS, 2) the adjusted association that would be derived by using Two-step *cis*-MR. The confounder-outcome association was estimated as the Wald ratio of association the non-cis genetic risk score for the confounder (GRS) with the outcome divided by the GRS-confounder association. The standard error was estimated as the standard error of the confounder (GRS) with the outcome divided by the absolute value of the GRS-confounder association.

*Performance measure:* The performance measure was the estimate of the casual effect, and the standard error in the estimate of the causal effect for the crude and Two-step cis-MR methods in settings in which multiple biasing pathways are adjusted for.

*Limitations of the product & dereference of coefficients methods*: The product & dereference of coefficients methods make strong assumptions, including:

A) no confounding in (1) the exposure-mediator association, (2) the mediator-outcome association, (3) the exposure-outcome association and (4) no confounders of the mediator-outcome association that are influenced by the exposure (note: in this setting, the variant is the exposure, and the mediator is the pleiotropic phenotype). (1), (2), and (3) are less likely in this setting because of the random inherence of genetic variants. (4) is unlikely because, by estimating the mediator-outcome association using MR, the major relevant confounder of the association would be residual population structure. Covariate adjustment is also less likely to introduce collider bias than in traditional applications because GWAS typically do not adjust for heritable phenotypes.

B) Regression dilution bias due to non-differential measurement error in the mediator will bias traditional implantations of this method. However, in the absence of Winner’s curse, MR is robust to non-differential measurement error so this assumption is less important.

C) It assumes no interaction/effect modification, although this is also typically an implicit assumption of Two-Sample MR.

*Linearity*: As with all summary data MR applications, we are assuming linear relationships.

*Model misspecification*: As with MVMR, TSCMR requires that we have correctly specified the pathway(s) in which the bias occurs.

*Estimation of the Standard Errors*: Both the bootstrapped SE and the propagation of error SE we calculate will be biased if the true standard error is not normally distributed.

*Instrumental variables assumptions*: as with any MR analysis, we assume that the assumptions of instrumental variables are valid for the MR analysis of the effect of the confounder on the outcome; that the only violation of the IV assumptions for the effect of the variant on the outcome is due to the confounder (i.e. that the variants are strongly associated with the exposure, only causes the outcome through the exposure or confounder, and is otherwise independent of the outcome conditional on the confounder); if two-sample MR is used, then we additionally require that the two-sample MR specific assumptions are valid. In addition, our method assumes that there is no bias in the variant-confounder association.

Supplementary Box 1: Non-exhaustive list of putative limitations of Two-Step cis-MR.

2^nd^ Pleiotropic phenotype

Genotype for 2^nd^ pleiotropic phenotype

Drug target genetic proxy

Drug target perturbation

Outcome

Pleiotropic phenotype

Genotype for pleiotropic phenotype

Supplementary Figure 1: DAG representing bias through pleiotropy or two variants in perfect LD, when there are two independent variables for adjustment.

2^nd^ Pleiotropic phenotype

Genotype for 2^nd^ pleiotropic phenotype

Drug target genetic proxy

Drug target perturbation

Outcome

Pleiotropic phenotype

Genotype for pleiotropic phenotype

Supplementary Figure 2: DAG representing bias through pleiotropy or two variants in perfect LD, when there are two non-independent variables for adjustment.

|  | Bias in Crude estimate | Bias in TSCMR Estimate | Crude Estimate SE | TSCMR PE SE | TSCMR BS SE |
| --- | --- | --- | --- | --- | --- |
| Sim. of Sup. Fig. 1 | 1.336 | -0.018 | 0.174 | 0.226 | 0.226 |
| Sim. of Sup. Fig. 2 | 0.406 | -0.365 | 0.174 | 0.248 | 0.248 |

Supplement Table 1: Results of simulation to explore the ability of 2SCMR to simultaneously adjust for multiple biasing pathways. Crude = MR estimates using the unadjusted variant-outcome association(s), TSCMR = MR estimates after adjusting the variant-outcome association(s) using TSCMR. SE = Standard error. PE = Propagation of Error. Supplementary Figure 1 is a setting in which both biasing paths are independent, Supplementary Figure 2 is a setting where the two paths are not independent. All estimates are the average of 100,000 receptions. Monte Carlo standard errors were all < 0.001, with the exception of bias in the both crude estimates (0.006 and 0.004 for the simulations of Supplementary Figure 1 and 2 respectively), and the adjusted estimate for the simulation of Supplementary Figure 2 (0.004)
